# A micro-costing analysis of tuberculosis care in England: a bottom-up evaluation of treatment and service delivery costs

**DOI:** 10.64898/2026.06.26.26356681

**Authors:** Srobana Ghosh, Sharon Cox, Esther Robinson, Martin Dedicoat, Y-Ling Chi

## Abstract

**Background:** In 2024 incidence of tuberculosis in England rose to 9.4 per 100,000, which is close to exceeding the low-incidence designation threshold. Addressing the rising incidence requires policy making to ensure sufficient staff, hospital resources and budgets are available to meet the increasing demand. However, costing of active pulmonary TB in the UK are limited and better clarity is needed on the clinical pathway of tuberculosis and the resources involved.

**Objective:** This study aims to estimate the costs of active pulmonary drug-sensitive tuberculosis care in England using a micro-costing approach.

**Method:** The analysis was performed from the perspective of the National Health Service (NHS), capturing direct medical costs only. The clinical pathway for different severities of tuberculosis care was defined through a review of the literature, clinical guidelines, and interviews with clinicians.

Costs were mainly drawn from the British National Formulary and the eMIT national database for drug costs, and the National Cost Collection (2021-22) for diagnostics, monitoring, nursing and hospitalisation alongside a desk-based review.

**Results:** Per-patient costs in 2021 ranged from approximately £2,000 for community-managed cases to over £50,000 for the most complex patients. An estimated 70% of patients cost between £4,971 and £7,307. The weighted average cost of treatment across all complexities was £8,125 per patient reflective of the proportion of cases at each severity. For the 4,423 patients in 2021, it is estimated that the costs of direct treatment were at least £36 million pounds, highlighting the significant financial implications of increasing tuberculosis.

**Conclusion:** The findings demonstrate that tuberculosis care imposes a substantial and highly variable cost burden on the NHS. Overall, this study provides cost estimates that can inform service planning, resource allocation, and future economic evaluations. Further research is needed on the costs of drug-resistant TB to support comprehensive TB control strategies.

**What is already known on this topic?:** Existing UK evidence on tuberculosis costs is limited with little detailed micro-costing of the full care pathway.

**What this study adds?:** This study provides granular, bottom-up estimates of tuberculosis care costs in England, demonstrating the drivers of costs and the substantial variation by disease complexity and severity.

**How this study might affect research, practice or policy?:** These findings can inform resource allocation and economic evaluations, supporting policies that prioritise early diagnosis and community-based care to reduce costs and healthcare burden.

## Introduction

Whilst the incidence of tuberculosis (TB) fell from its peak in 2012 to its lowest level in 2020, cases have since been rising each year and have reached 5,490 new notifications in 2024(1). This is a 13% increase on the 4,831 cases in 2023, which was a 11% increase on the cases in 2022(2). The TB notification rate in England is 9.4 per 100,000, which is close to exceeding the World Health Organisation’s designated rate of 10 per 100,000 for a low incidence country(1).

There is an additional equity component as tuberculosis disproportionately affects the poorest and most vulnerable members of society. Additionally, there is a health security risk from tuberculosis as 2%(1) of cases are drug resistant or potentially multidrug resistant (MDR), adding to concerns of growing anti-microbial resistance in England(3).

Addressing the uptick in tuberculosis incidence requires policy making and planning to ensure sufficient staff, hospital resources and budget are available to meet the increasing demand. The United Kingdom Health Security Agency (UKHSA) and National Health Service England (NHSE) are overseeing the national action plan for TB. Meanwhile, clinics and public institutions such as prisons are enhancing their tuberculosis capabilities(4-7).

Costs of tuberculosis care are needed to determine budgets, allocate resources and develop feasible strategies to address the escalating rate of TB. However, there is limited recent evidence on the costs of active pulmonary tuberculosis care for the general population in England (4, 5, 7-10). Better clarity is also needed on the clinical pathway of tuberculosis and the resources involved.

Micro-costing, or bottom-up costing, provides a detailed approach to estimating healthcare costs by identifying and valuing individual resource components. This method is particularly useful for complex conditions such as TB, where care pathways involve multiple stages and resource inputs.

This study aims to fill this gap by providing an up to date and validated estimate of the cost of TB care in England using a micro-costing approach. Specifically, we quantify the per-patient cost of active pulmonary drug-sensitive TB, whilst identifying key cost drivers across the care pathway.

## Methods

### Study design and perspective

A bottom-up micro-costing analysis was conducted to estimate the costs of active drug-sensitive pulmonary tuberculosis care. The analysis was performed from the perspective of the NHS, capturing direct medical costs only.

The analysis included all tuberculosis notifications in England in 2021, categorised by severity. The severity categories were taken from the National Cost Collection (NCC)(11) Hospital Resource Group (HRG)(12) complications/comorbidity (CC) scores which categorises and reflects increasing levels of care complexity based on procedural requirements and comorbidity burden. Severity categories include:

- Complex Tuberculosis
- TB with interventions
- TB (without interventions, CC 7+)
- TB (without interventions, CC 3-6)
- TB (without interventions, CC 0-2)
- Community managed TB (no hospital admission)

Complex tuberculosis patients are those who require a lengthy admission (over 28 days) usually due to social problems such as homelessness or having TB that is difficult to treat, including MDR, and therefore have an enhanced tariff(13). The other HRG codes reflect a shorter length of stay with declining severity defined by the level of interventions and complications as reflected in the CC score(13). Community managed TB assumed that no hospital admission was required.

Whilst the HRG for complex patients includes the hospital care for MDR patients, a full costing of MDR or extensively drug-resistant (XDR) patients could not be completed due to a lack of data. A speculative scenario calculating the average costs of tuberculosis per patient including the costs of MDR was conducted using estimates from the literature as an illustrative example.

### Care pathway and resource identification

We defined the clinical pathway for different severities of tuberculosis through a review of the literature, evaluating NICE guidelines, and interviews with clinicians. Clinician interviews were also used to verify and validate our assumptions of the clinical pathway and develop the finalised version.

The TB care pathway was divided into four main components:

- Diagnostics and initial assessment
  º Laboratory tests (e.g. sputum microscopy, smear and culture, whole genome sequencing for drug sensitivity)
  º Imaging (e.g. chest radiography, computerised tomography scans)
- Pharmacological treatment
  º First-line antibiotics for drug sensitive tuberculosis
- Monitoring and follow-up
  º Initial appointment to set the treatment protocol and conduct contact tracing
  º Routine clinical reviews throughout the six months of treatment, including follow up tests
  ▪ Additional support face to face (directly observed therapy (DOTs)), or via video call (video observed therapy (VOTs))
  º Discharge appointment
- Hospitalisation costs
  º Inpatient admissions by severity

Resource use for each component was estimated based on standard treatment as per clinical guidelines, expert input, or aggregated data. Variations in care could not be fully costed, which was a limitation of the analysis.

## Data collection

Patient numbers were taken from the ‘TB in England 2021-22’ report(14), whilst the distribution of patients across the different severity groups was informed by the National Cost Collection’s tuberculosis HRGs(11).

The NCC for the NHS comprises aggregated costs (the average unit cost of providing defined services to NHS patients in England) and patient-level costs based on the specific interactions a patient has, and the events related to their healthcare activity(11). They are a key source of information about the cost of NHS services. HRGs are standard groupings of clinically similar treatments, procedures, and diagnoses that consume similar levels of NHS healthcare resources(12).

Costs were mainly drawn from the British National Formulary(15) and the eMIT national database(16) for drug costs, and the National Cost Collection (2021-22)(11) for diagnostics, monitoring, nursing and hospitalisation alongside a desk-based review. A small proportion are likely to require more invasive procedures to diagnose their TB and it is assumed this is included in the HRGs. Drug costs were based on the assumption of patients receiving the standard fixed dose combination, although there may be more variation in clinical practice. Drugs used to mitigate the side effects of TB medications and reduce the symptoms of TB have not been costed due to the inconsistency of their use in the clinical pathway.

## Analysis

The total cost of tuberculosis care per patient by severity was calculated by summing costs across all components of care. Costs were also disaggregated to determine the contribution of each component.

Total costs to the health system were calculated by multiplying the costs of care by the total population, accounting for severity of illness. The average cost of care is a weighted average of all disease severities.

Scenario analyses were conducted deterministically by modifying the key cost parameters of drug prices to compare list prices against generic.

Costs are reported in 2021 GBP £.

## Results

There were 4,423 total TB notifications in 2021(14). By severity there were 3% (147) complex patients, 16% (697) had treatment that required intervention, 27% (1,187) had TB which did not require an intervention with a CC score of 7+, 24% (1,052) had TB(CC 3-6) which did not require an intervention, 21% (909) had TB (CC 0-2) that did not require an intervention and 10% (431) had TB that could be managed in the community without hospitalisation(14)(Table 1).

**Table 1:**
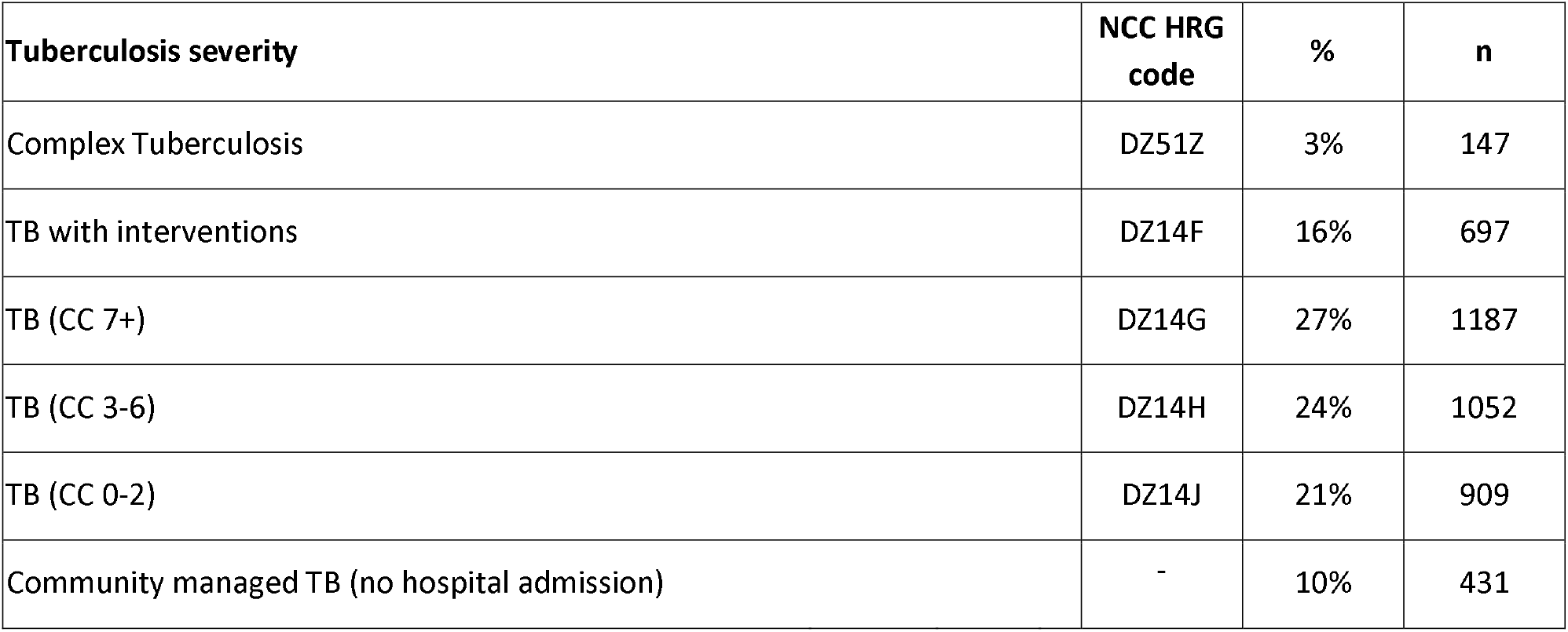
Tuberculosis patients by severity.

**Table 2:** Tuberculosis diagnostic costs.

| Item | Cost | Year | 2021 cost | Source | Complex TB | TB with interventions | TB (CC 7+) | TB (CC 3-6) | TB (CC 0-2) | No hospital admission |
| --- | --- | --- | --- | --- | --- | --- | --- | --- | --- | --- |
| Chest x-ray | £41 | 2022 | £39 | National Cost Collection Year 2021/22 Plain film outpatient | 3 | 3 | 3 | 3 | 3 | 3 |
| Computerised Tomography Scan of One Area | £90 | 2022 | £86 | National Cost Collection Year 2021/22 RD20A | 1 | 1 | - | - | - | - |
| Laboratory cost: smear and culture | £35 | 2022 | £33 | Internal UKHSA estimate | 1 | 1 | 1 | 1 | 1 | 1 |
| Sputum test | £7 | 2011 | £9 | Jit et al 2011 | 3 | 3 | 3 | 3 | 3 | 3 |
| Drug sensitivity test to test for resistance (WGS) | £116 | 2022 | £111 | Internal UKHSA estimate | 1 | 1 | 1 | 1 | 1 | 1 |
| **Total costs of diagnosis by severity** |  |  |  |  | **£373** | **£373** | **£288** | **£288** | **£288** | **£288** |

**Table 3:** Antibiotic dosage and pack size.

| Drug | Dose | Tablets in pack | Mg/ tablet | Source |
| --- | --- | --- | --- | --- |
| Isoniazid | 300 mg daily for 6 months | 28 | 100 | https://bnf.nice.org.uk/drugs/isoniazid/medicinal-forms/<br>https://www.gov.uk/government/publications/drugs-and-pharmaceutical-electronic-market-information-emit |
| Rifampicin | 600 mg once daily for 6 months (initial and continuation phases) | 60 | 300 | https://bnf.nice.org.uk/drugs/rifampicin/medicinal-forms/ |
|  |  |  |  | https://www.gov.uk/government/publications/drugs-and-pharmaceutical-electronic-market-information-emit |
| Pyrazinamide | 2 g once daily for 2 months (initial phase). | 30 | 500 | https://bnf.nice.org.uk/drugs/pyrazinamide/medicinal-forms/ |
| Ethambutol | 15 mg/kg once daily for 2 months.<br>Assuming an average of men and women.<br>In 2021, the mean weights of men and women based on adjustments to self-reported weight were 85.1kg and 71.8kg, respectively. https://digital.nhs.uk/data-and-information/publications/statistical/health-survey-for-england/2021/part-4-trends | 56 | 100 | https://bnf.nice.org.uk/drugs/ethambutol-hydrochloride/ |

**Table 4:** Antibiotic costs.

| Drug(15) | Dose (mg) | Dose/day | Total days | Mg / pack | Packs / dose | Cost per pack(16) | Cost per day | Total cost of treatment |
| --- | --- | --- | --- | --- | --- | --- | --- | --- |
| Isoniazid | 300 | 1 | 180 | 2800 | 0.11 | £8.26 | £0.89 | £159 |
| Rifampicin | 600 | 1 | 180 | 18000 | 0.03 | £25.19 | £0.84 | £151 |
| Pyrazinamide | 2000 | 1 | 60 | 15000 | 0.13 | £11.24 | £1.50 | £90 |
| Ethambutol | 1176.75 | 1 | 60 | 5600 | 0.21 | £5.09 | £1.07 | £64 |
| **Total** |  |  |  |  |  |  |  | **£465** |

**Table 5:** Total costs of tuberculosis care by severity.

| Total patients by severity | Percentage of total cases | Number | Screening | Drug costs | Routine monitoring | Hospital costs | DOTs | VOTs | Total costs per patient | Total costs for all pts by severity |
| --- | --- | --- | --- | --- | --- | --- | --- | --- | --- | --- |
| Complex Tuberculosis | 3% | 147 | £373 | £536 | £1,345 | £28,296 | £19,056 | £5,007 | £54,614 | £8,028,199 |
| TB with interventions | 16% | 697 | £373 | £536 | £1,345 | £7,360 | £1,045 | £275 | £10,934 | £7,621,175 |
| TB (CC 7+) | 27% | 1187 | £288 | £536 | £1,345 | £4,649 | £388 | £102 | £7,307 | £8,673,539 |
| TB (CC 3-6) | 24% | 1052 | £288 | £536 | £1,345 | £3,294 | £310 | £81 | £5,854 | £6,158,328 |
| TB (CC 0-2) | 21% | 909 | £288 | £536 | £1,345 | £2,464 | £268 | £71 | £4,971 | £4,518,962 |
| Community managed TB (no hospital admission) | 10% | 431 | £288 | £536 | £1,345 | £0 | £0 | £0 | £2,168 | £934,620 |
| Weighted average cost of active TB per patient | £8,125 |  |  |  |  |  |  |  |  |  |
| Costs for all patients | £36 m |  |  |  |  |  |  |  |  |  |

For a diagnosis of active pulmonary TB, all patients were expected to have three chest x-rays (£39 each(11)), one laboratory smear and culture (£33(17)), three sputum tests (£9 each(5)) and one whole genomic sequencing drug sensitivity test (£111(17)). Total costs of diagnostics and imaging were £288. The most complex patients (complex and treatment with interventions) had an additional computerised tomography scan costing £86(11), bringing their total diagnostic and imaging costs to £373.

The dose for a standard regimen for active pulmonary TB includes isoniazid and rifampicin for six months with pyrazinamide and ethambutol for three months. It was assumed that each complexity has the same standard cycle of antibiotics. Total costs of antibiotics for the entire regimen were £465 using the eMIT database(16). A scenario using list prices from the BNF found that drug costs would have been £2,180.

Clinician interviews informed that every patient is entitled to 6 routine nurse appointments across their care pathway for the diagnostic work up, start of treatment (including contact tracing), two-week review, one month review, five-month review and at the end of treatment. These appointments were costed at £224 using cost code DZ42Z(11), totalling £1,345. It is assumed the HRGs include the costs of follow up tests, if they were required. Some patients require additional support in the form of DOTs and VOTs costing £131 and £52 respectively(11). The use of DOTs and VOTs were allocated to patients requiring hospital care in the same proportion as hospitalisation costs per severity.

Hospitalisation costs were taken from the National Cost Collection 2021/22(11) using cost codes DZ51Z (complex), DZ14F (treatment with interventions), and for treatment without interventions DZ14G (CC 7+), DZ14H (CC 3-6), DZ14J (CC 0-2) respectively costing £28,296, £7,360, £4,649, £3,294 and £2,464.

Total costs of treatment for active pulmonary TB ranged widely with severity ranging from £2,168 for the simplest cases to at least £54,614 for the most complex. Approximately 70% of patients cost between £4,971 and £7,307. The weighted average cost of treatment across all complexities was £8,125 reflective of the proportion of cases at each severity.

For the 4,423 patients in 2021, it is estimated that the costs of direct treatment were at least £36 million pounds.

The main driver of costs appears to be hospital costs, particularly the severity of tuberculosis. Drug costs are also a driving factor. These costs used the eMIT prices that should reflect commercial discounts and generic prices. Had the study used list prices for drugs, the average cost would be closer to £10,000 per patient weighted across severity in 2021 prices.

If 2% of TB notifications were MDR or XDR, there would have been 88 such patients. Using an estimate from the literature for the costs of MDR care of £100,000 per person would have cost £8.8 million. This would bring the total costs of all TB notifications to £41 million, with an average cost of patients as £9,177.

## Discussion

This study provides a detailed micro-costing analysis of the cost of active pulmonary tuberculosis care in England, addressing a key evidence gap in the current literature. The results demonstrate that TB treatment imposes a substantial financial burden on the NHS, costing on average approximately £8,000 per case although the costs can range from £2,000 to £54,000, ultimately costing £36 million across all patients in 2021.

As cases escalate, the total costs of TB care will escalate also. In 2024 there were 24% more notifications than in 2021, and likely would have costed £54 million. Beyond escalating treatment costs, the increase in TB notifications entails that there is a commensurate higher need for nurses, TB units, scans, radiographers and antibiotics in an already stretched system. Severity of tuberculosis can either be social or clinical, with some complex patients requiring daily appointments to manage their care. The National Action Plan identified London and Leicester as areas with the highest number of notifications(1, 18), but other areas are also seeing an increase in cases and may not have sufficient resources to provide care. Scaling up and distributing services is complex and requires considerable resources including training, infrastructure and equipment which may be lacking particularly in certain regions. Tuberculosis in settings such as prisons can also add to resource constraints.

The analysis focused on drug sensitive tuberculosis, however 2%(1) of cases are drug resistant and some are extremely drug resistant which has the potential to drive costs even higher with alarming consequences for national antimicrobial resistance resilience. The literature is inconsistent on the total costs of MDR and XDR care with considerably high upper estimates(9, 10). As cases increase, it is unknown what proportion could be MDR or XDR and the ensuing implications.

The analysis has demonstrated that TB care is resource-intensive and highly heterogenous across disease severities and drug resistance. The results could be used to inform economic models exploring the impacts of tuberculosis. Costs are driven by complexity, care setting and uncertainty in unit costs. The costing assumed that patients would receive the standard fixed dose drug regimen, and standard diagnostics and monitoring, however in clinical practice there would be more variability. A small proportion are likely to require more invasive procedures to diagnose their TB such as an ultrasound, MRI or diagnostic bronchoscopy but these were assumed to be included in the more complex patient’s HRG codes. Patients might require additional support such as but include proton pump inhibitors, antiemetics, analgesics, vitamins, antihistamines and corticosteroids, but these could not be costed due to variations in clinical practice. More work is needed to sufficiently capture the costs of complex care, particularly for MDR and XDR and the costs of providing care in settings outside of the hospital. Such evidence could inform investments for screening and developing TB programmes to encourage earlier diagnosis so that more patients with lower severity can be treated through community management.

## Limitations

### This study has several limitations

Resource use was based on an average estimate, which may not reflect all costs and the real-world variation in care and costs. The care pathway in the analysis includes the standard course of antibiotics and associated nursing and hospitalisation costs but does not account for variations in care or post-TB care should patients fail to respond to treatment or relapse. Here the costs are likely to increase.

Unit costs may vary across NHS settings, particularly for drug costs if commercial drug discounts are not generalisable. Should new antibiotics become available, the costs are likely to escalate higher. A conservative approach was taken to costing, and so costs could potentially be higher.

The analysis only costed direct hospital costs to reflect the costs to the NHS and did not include costs of primary care, social, indirect or patient-incurred costs. The true costs of care could therefore be higher.

## Conclusion

This analysis is a micro-costing study of active pulmonary tuberculosis care in England, addressing a key gap in the current economic evidence. The findings demonstrate that TB care imposes a substantial and highly variable cost burden on the NHS, with per-patient costs in 2021 ranging from approximately £2,000 for community-managed cases to over £50,000 for the most complex patients. Approximately 70% of patients cost between £4,971 and £7,307. The weighted average cost of treatment across all complexities was £8,125 per patient in 2021 (£10,351 in 2026) reflective of the proportion of cases at each severity. For the 4,423 patients in 2021, it is estimated that the costs of direct treatment were at least £36 million pounds, highlighting the significant financial implications of TB at a population level.

Per patient costs are driven primarily by social or clinical complexity, with higher need patients accounting for a disproportionate share of total costs. Whilst most patients are associated with moderate cost ranges, a small proportion of complex cases generate substantial costs particularly where prolonged admissions or intensive management are required. Other drivers of cost are increases in drug costs, patient relapse, and the potential increase of MDR or XDR TB. As TB incidence continues to rise in England, these findings suggest that the associated costs will also rise, as well as healthcare resource demands placing additional pressure on the NHS. These all underscore the importance of prevention, early detection, and effective treatment adherence.

Overall, this study provides cost estimates that can inform service planning, resource allocation, and future economic evaluations of TB interventions in the UK. Further research is needed to better characterise the costs of drug-resistant TB, long-term outcomes, and care delivered outside hospital settings to support comprehensive TB control strategies.

## Data Availability

All data produced in the present study are available upon reasonable request to the authors

## Declarations

### Contributors

Concept and design: SG

Acquisition of data: SG, ER, SC

Analysis and interpretation of data: SG

Manuscript first draft: SG

All authors reviewed and edited the manuscript.

Obtaining funding: ER, SC, YLC

Supervision: ER, SC, MD, YLC Guarantor: YLC

### Patient and public involvement

Patients and/or the public were not involved in the design, or conduct, or reporting, or dissemination plans of this research

### Provenance and peer review

Not commissioned; externally peer reviewed.

### Data availability statement

Data are available upon reasonable request.

### Patient consent for publication

Not applicable.

### Ethics approval

Not applicable.

